# Don’t wait, re-escalate: delayed action results in longer duration of COVID-19 restrictions

**DOI:** 10.1101/2020.11.04.20226316

**Authors:** Amy Hurford, James Watmough

## Abstract

Non-pharmaceutical public health interventions have significant economic and social costs, and minimizing their duration is paramont. Assuming that interventions are sufficient to reduce infection prevalence, we use a simple linear SIR model with case importation to determine the relationship between the timing of restrictions, duration of measures necessary to return the incidence to a set point, and the final size of the outbreak. The predictions of our linear SIR model agree well with COVID-19 data from Atlantic Canada, and are consistent with the predictions of more complex deterministic COVID-19 models. We conclude that earlier re-escalation of restrictions results in shorter disruptions, smaller outbreaks, and consequently, lower economic and social costs. Our key message is succinctly summarized as ‘don’t wait, re-escalate’ since delaying re-escalation of restrictions results in not only more infections, but also longer periods of restrictions.

## 1 Introduction

Non-pharmaceutical public health interventions are an essential tool during a pandemic. Indeed, we can expect they will often be the only available tools at the start of a pandemic when little is known about a novel pathogen. However, these interventions have significant economic and social costs, and minimizing the duration and impact of interventions is paramount. In Canada, and in particular in Atlantic Canada and the Territories, contact tracing and testing combined with travel restrictions, quarantine of incoming travellers, and social distancing, including business closures and limits on larger gatherings are the main tools in use during the current severe acute respiratory syndrome coronavirus 2 (SARS-CoV-2) pandemic. These have proven very effective in the smaller centres. At the time of writing border restrictions and traveller quarantine in conjunction with testing and contact tracing have allowed most businesses and social activity to resume a new, albeit slightly restricted, normal.

Current modelling efforts to understand the relative efficiency of the interventions include a full range of of models from simple phenomenological forecasts [12] to complex network and agent-based models explicitly including households and healthcare systems [1, 6, 15]. While these details are necessary to predict healthcare needs and monitor capacity, we argue that the strategy for timing of implementing non-pharmaceutical community interventions can be based on much simpler linear deterministic models.

Specifically, we use a simple linear SIR model with case importation to determine the relationship between the timing of restrictions, duration of measures necessary to return the incidence to a set point, and the final size of the outbreak. We conclude from our analysis that delaying re-escalation of restrictions leads to increased duration of control measures and larger outbreaks. Conversely, earlier re-escalation results in shorter disruptions, smaller outbreaks, and consequently, lower economic and social costs.

## 2 Linear SIR model

Consider a linear Susceptible-Infected-Recovered [8] model,

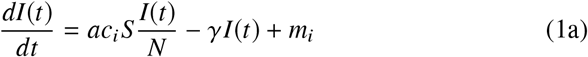

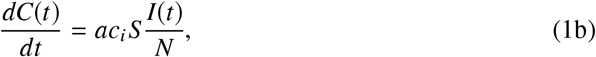

where *I*(*t*) is the number of infected individuals, and *C*(*t*) is the cumulative cases at time, *t*. The contact rate, *c*_*i*_, is the rate that a susceptible individual contacts another individual in the population, 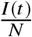 is the probability that the contacted individual is infected, where *N* is the total population size, and *a* is the probability of infection given a contact. The rate that infected individuals recover is *γ* per day and the rate that cases are imported is *m*_*i*_ cases per day. The appropriate initial values are *I*(*0*) = *I*_0_ and *C*(0) = 0.

The number of susceptible individuals in the population is *S*, and as an approximation, *S* is assumed to be unchanging. This assumption is justified when the number of susceptible individuals is very large, and only a small fraction of these susceptible individuals have been infected during the study period. This is the case in many regions, for example, in Canada, as of October 29, 2020, there have been 228,542 clinical coronavirus disease 2019 (COVID-19) cases [9], which represents 0.6% of the Canadian population [10]. If 40% of cases are asymptomatic, then only around 1% of the Canadian population has been infected to date.

We consider the epidemiological dynamics subject to restrictions that affect the contact rate and the importation rate, and are implemented at *t* = *t*_1_. Let *c*_*i*_ and *m*_*i*_ denote the contact and importation rates where *i* = 1 indicates prior to restrictions (0 ≤ *t* ≤ *t*_1_), and *i* = 2 indicates after restrictions (*t*_1_ ≤ *t* ≤ *t*_2_). To simplify the notation, let *β*_*i*_ = *ac*_*i*_/ *N* and *λ*_*i*_ = *β*_*i*_*S* −*γ*.

We consider a restricted parameter space, such that the number of infected individuals increases until the restrictions are implemented, and then decreases. This scenario represents the epidemiological dynamics for outbreaks that have subsequently been eradicated, for example outbreaks in New Zealand, Taiwan [17], and the provinces of Atlantic Canada [9]. The parameter space that we consider is such that prior to restrictions the outbreak grows. There are two parameter combinations that allow for this possibility: (1) importations occur, *m*_1_ > 0, and *λ*_1_ > −*m*_1_/*I*_0_; or (2) if importations do not occur, *m*_1_ = 0, then *λ*_1_ > 0. After restrictions, we assume that *λ*_2_ < 0 and *m*_2_ = 0, such that the outbreak dissipates, and infection is eventually eradicated.

Under these assumptions, the outbreak exponentially increases and then exponentially decreases, and we can solve Equation 1a, such that,

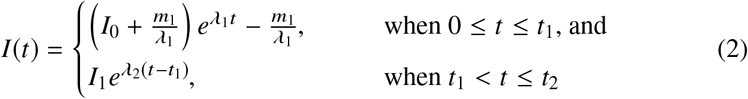

where *I*_1_ is the number of infected individuals when restrictions are implemented:

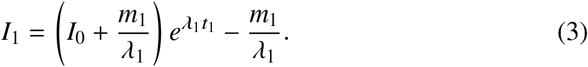

Note that after the restrictions are enacted (*t* > *t*_1_), the number of infections, *I*(*t*), is decreasing (since *λ*_2_ < 0), and when the number of infected individuals decreases below a value *I*_2_, we assume that the restrictions might be lifted, and then the duration of the restrictions is calculated as

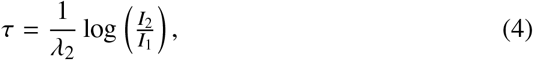

where we have used log to denote the natural logarithm. If *λ*_2_ < 0 < *λ*_1_, or if *λ*_2_ < 0 and *λ*_1_ > −*m*_1_/*I*_0_, the duration of restrictions, τ, is increasing with respect to the timing of restrictions, *t*_1_:

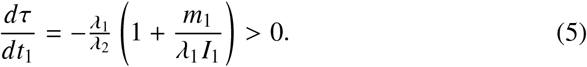

Hence, delaying the implementation of the restrictions results in a longer period that the restrictions must be in place to reduce the active number of cases, *I*(*t*), below the target *I*_2_. If *λ*_2_ < *λ*_1_ < −*m*_1_ /*I*_1_, then τ is decreasing with respect to *t*_1_. Since in this last scenario, the number of active cases, *I*(*t*) is decreasing for 0 < *t* < *t*_1_, it is not likely a scenario that would trigger re-escalation, and we do not consider it further.

Next, we calculate the total number of infections (also referred to as the final size [4]). The cumulative number of infections at the time when the restrictions are implemented is,

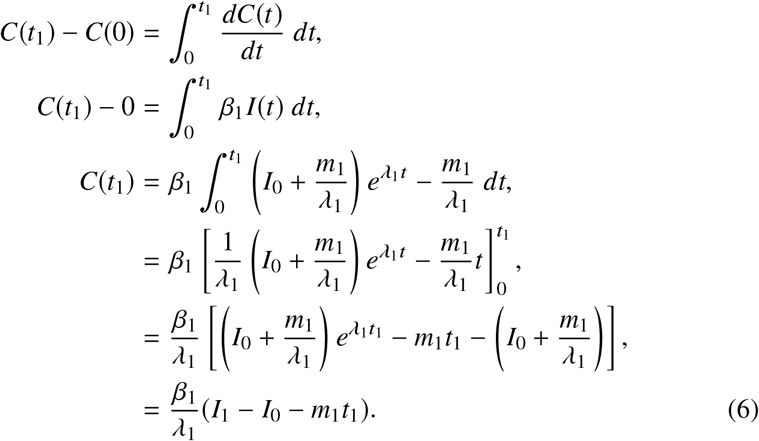

The total number of cases occurring after restrictions, which we denote *C*(*t*_*2*_), is found by a simple integration:

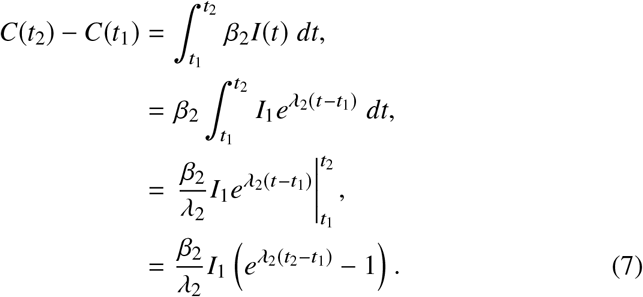

Therefore, adding Equations 6 and 7, the total number of infections in the outbreak is

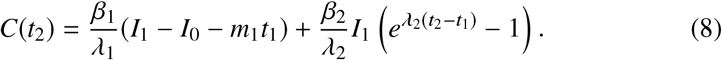

Returning to Equation 7, and rearranging Equation 6 such that 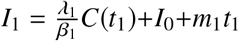, we have that,

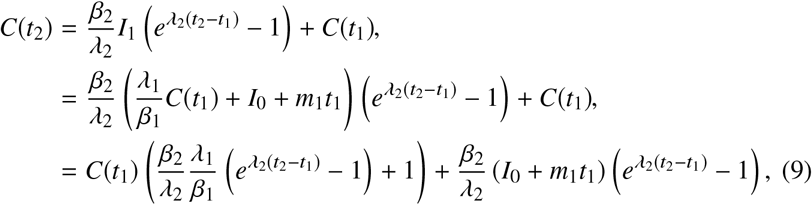

such that the total number of cases in the outbreak is a linearly increasing function oof the total number of cases up until the restrictions are enacted, with a slope of 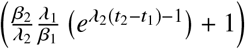.

## 3 Comparing the linear SIR model predictions to data

To test the predictions of the linear SIR model, we fit Equation 2 to data describing the number of active infections in Newfoundland and Labrador (NL) and New Brunswick (NB) for the March-May 2020 outbreaks that occurred in these Canadian provinces (Figure 3; where these data are from [7]). We defined *t* = 0 as the day that the number of active cases first exceeded 10. We then use the estimated values for *λ*_1_, *λ*_2_ and *m*_1_ to predict the number of active cases at the peak of the outbreak (Equation 3), the required duration of predictions (Equation 4), and the total number of cases in the outbreak (Equation 8 and Table 1). We assumed that restrictions were enacted on the day, *t*_1_, corresponding to the peak number of active infections in the data. Fitting was performed using maximum likelihood and assuming normally distributed residuals.

**Table 1.**
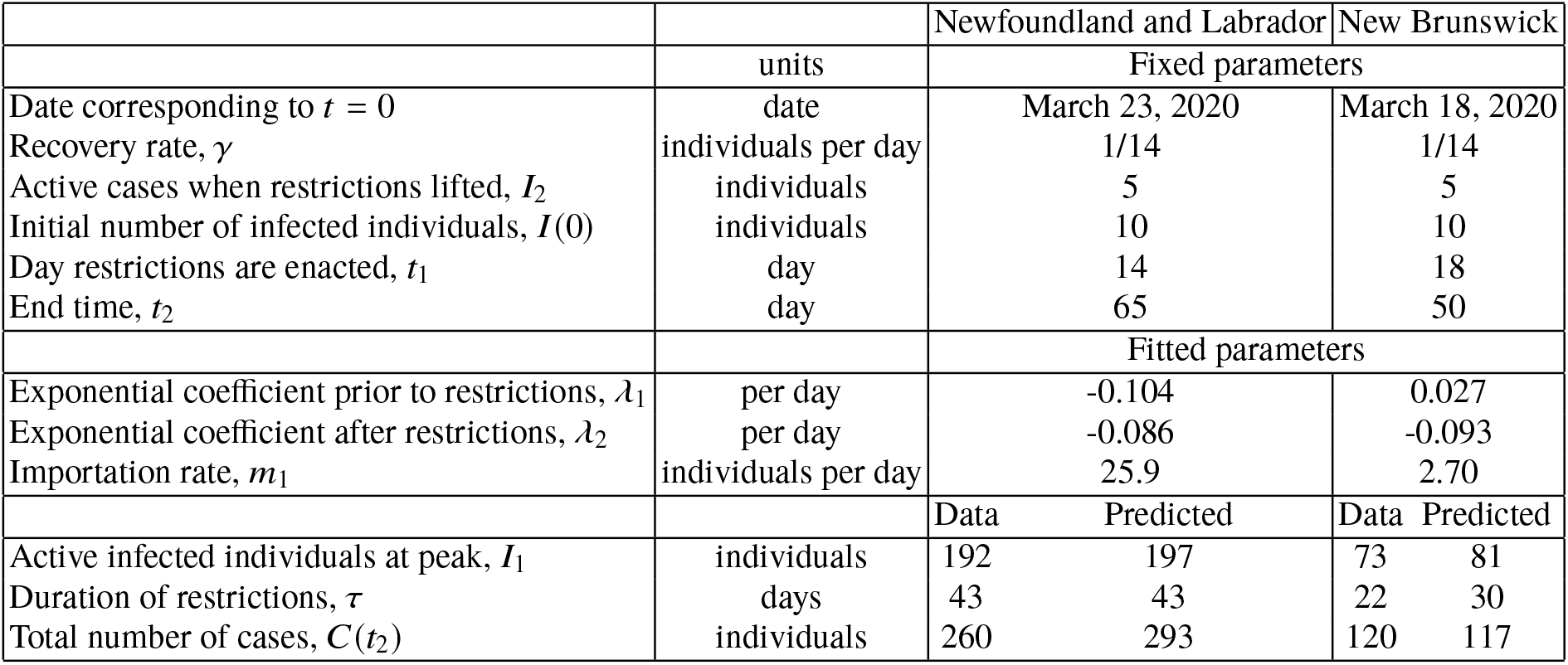
Parameter estimates and validation of the linear SIR model fitted to active case data (see Figure 3)

**Table 2.** Parameters and initial values for the linear SIR and the COVID-19 model.

| COVID-19 model |  |  |  |  |
| --- | --- | --- | --- | --- |
| Description | symbol | units | value | reference |
| Transmission rate prior to restrictions | $\beta_1$ | per time | 0.571 | to achieve 7 day doubling time |
| Transmission rate after to restrictions | $\beta_2$ | per time | 0.185 | to achieve 10 day halving time |
| Infectiousness of clinical relative to pre-clinical cases | $b_C$ | unitless | 0.5 | [11, 14] |
| Infectiousness of asymptomatic relative to pre-clinical cases | $b_A$ | unitless | 0.5 | [11, 14] |
| Rate of become infectious after exposure | $\delta_L$ | per time | 1/3 | [11, 14] |
| Rate of developing symptoms after becoming infectious | $\delta_P$ | per time | 1/2.1 | [11, 14] |
| Rate of recovery for a clinical infection | $\delta_C$ | individuals per time | 1/2.9 | [11, 14] |
| Rate of recovery for an asymptomatic infection | $\delta_A$ | individuals per time | 1/5 | [11, 14] |
| Proportion of cases that are symptomatic | $r$ | unitless | 0.6 | estimates vary, but see [11] |
| Initial susceptible | $S(0)$ | individuals | 99,990 | |
| Initial with preclinical infection | $I_P(0)$ | individuals | 1.5 | |
| Initial with clinical infection | $I_C(0)$ | individuals | 2.5 | |
| Initial with asymptomatic infection | $I_A(0)$ | individuals | 3.0 | |
| Initial latently infected | $L(0)$ | individuals | 3.0 | |
| Linear SIR model |  |  |  |  |
| Description | symbol | units | value | reference |
| Transmission rate prior to restrictions | $\beta_1 S$ | per time | 0.224 | to achieve 7 day doubling time |
| Transmission rate after to restrictions | $\beta_2 S$ | per time | 0.057 | to achieve 10 day halving time |
| Rate of importations prior to restrictions | $m_1$ | cases per time | 0 | to be consistent with the COVID-19 model |
| Rate of recovery | $\gamma$ | per time | 1/8 | $= 1/\delta_L + 1/\delta_A$ to be consistent with the COVID-19 model |
| Initial infected | $I(0)$ | individuals | 10 | |
| Initial cumulative infections | $C(0)$ | individuals | 0 | |
| Both models |  |  |  |  |
| Description | symbol | units | value | reference |
| Total population size | individuals | $N$ | 100,000 | |
| Number of active cases when restrictions end | cases | $I_2$ | 10 | |
| Day when restrictions are enacted | day | $t_1$ | 1 to 45 days | |

**Fig. 1.**
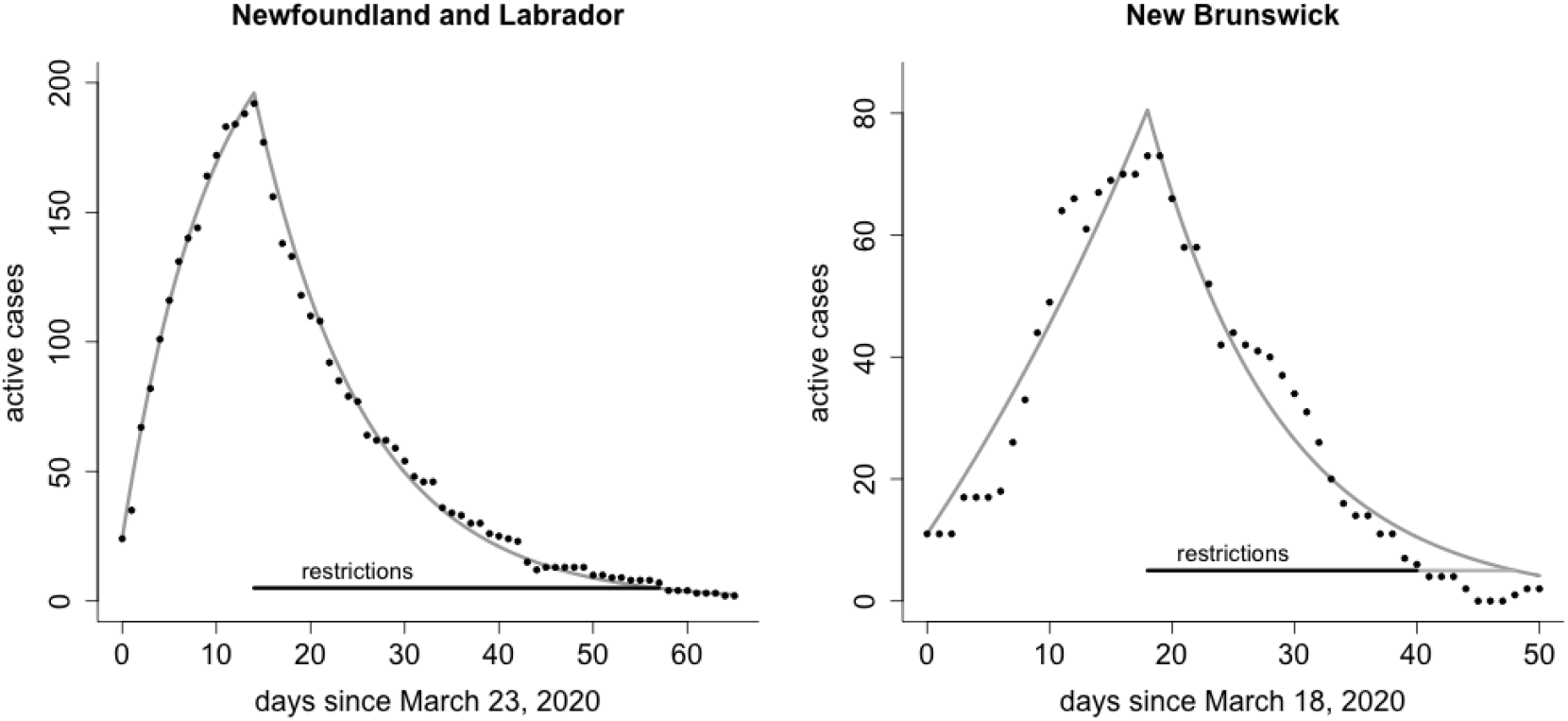
The linear SIR model fitted to data describing the active number of COVID-19 cases in Newfoundland and Labrador and New Brunswick from March-May, 2020. Equation 1a, describing the number of active infections for the linear SIR model (grey line), is fit to data describing the number of active COVID-19 cases (black dots). We assume that restrictions are enacted at the time of the peak number of active infections in the data and remain until there are fewer than 5 active cases (black horizontal line). The duration of restrictions, τ, is predicted by the linear SIR model (Equation 4; grey horizontal line). The fitted model parameters and the comparison of the model predictions with the data are reported in Table 1.

**Fig. 2.**
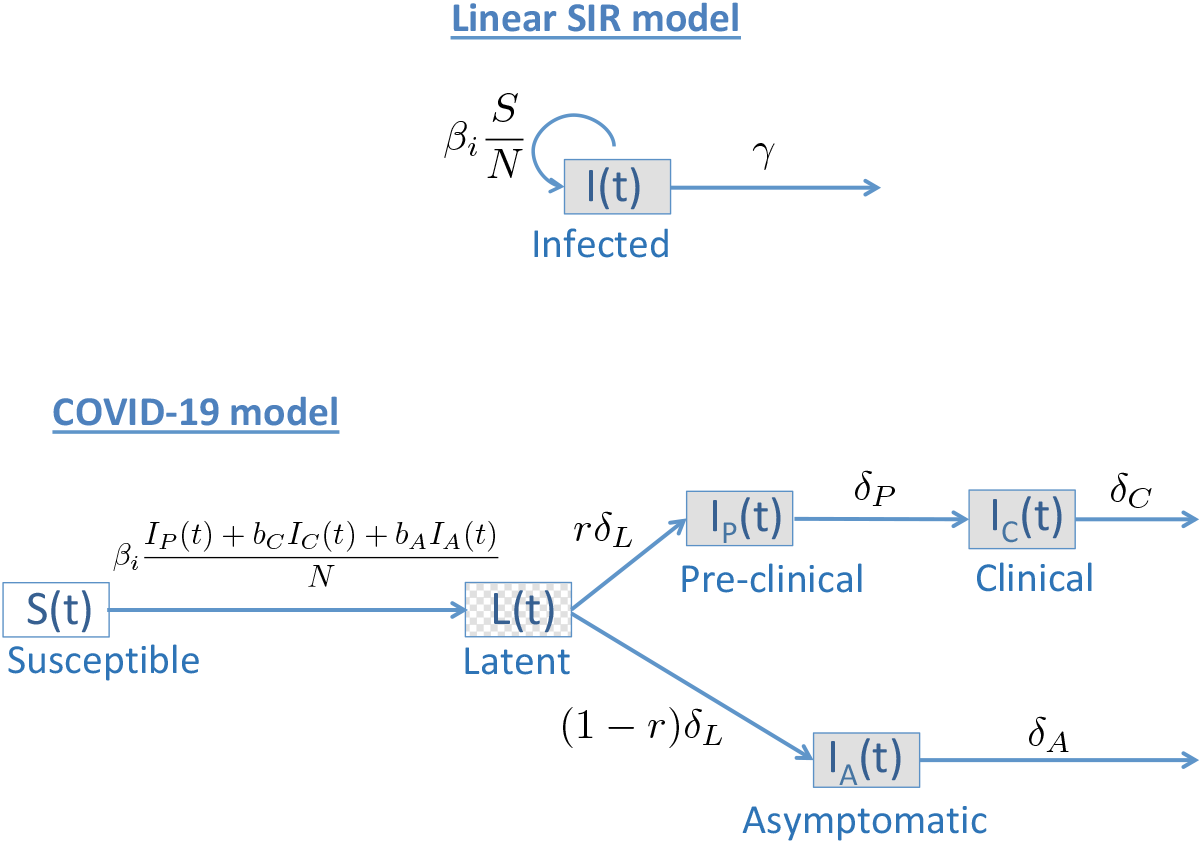
Diagram comparing the Linear SIR model and the COVID-19 model. The linear SIR model has just one dynamic state, the number of infected individuals, *I*(*t*), and the number of susceptible individuals, *S*, is assumed to be unchanging. The COVID-19 model (Equations 10) has five dynamic states: susceptible individuals, *S*(*t*), latently infected individuals, *L*(*t*), and individuals with: pre-clinical infections (not yet showing symptoms), *I*_*P*_(*t*); clinical infections (showing symptoms), *I*_*C*_(*t*); and asymptomatic infections, *I*_*A*_(*t*). States representing infected individuals have some grey fill, infectious states have solid grey fill, and since latently infected individuals are not infectious this compartment has a checkered grey fill. All model parameters are defined in Table 4.

**Fig. 3.**
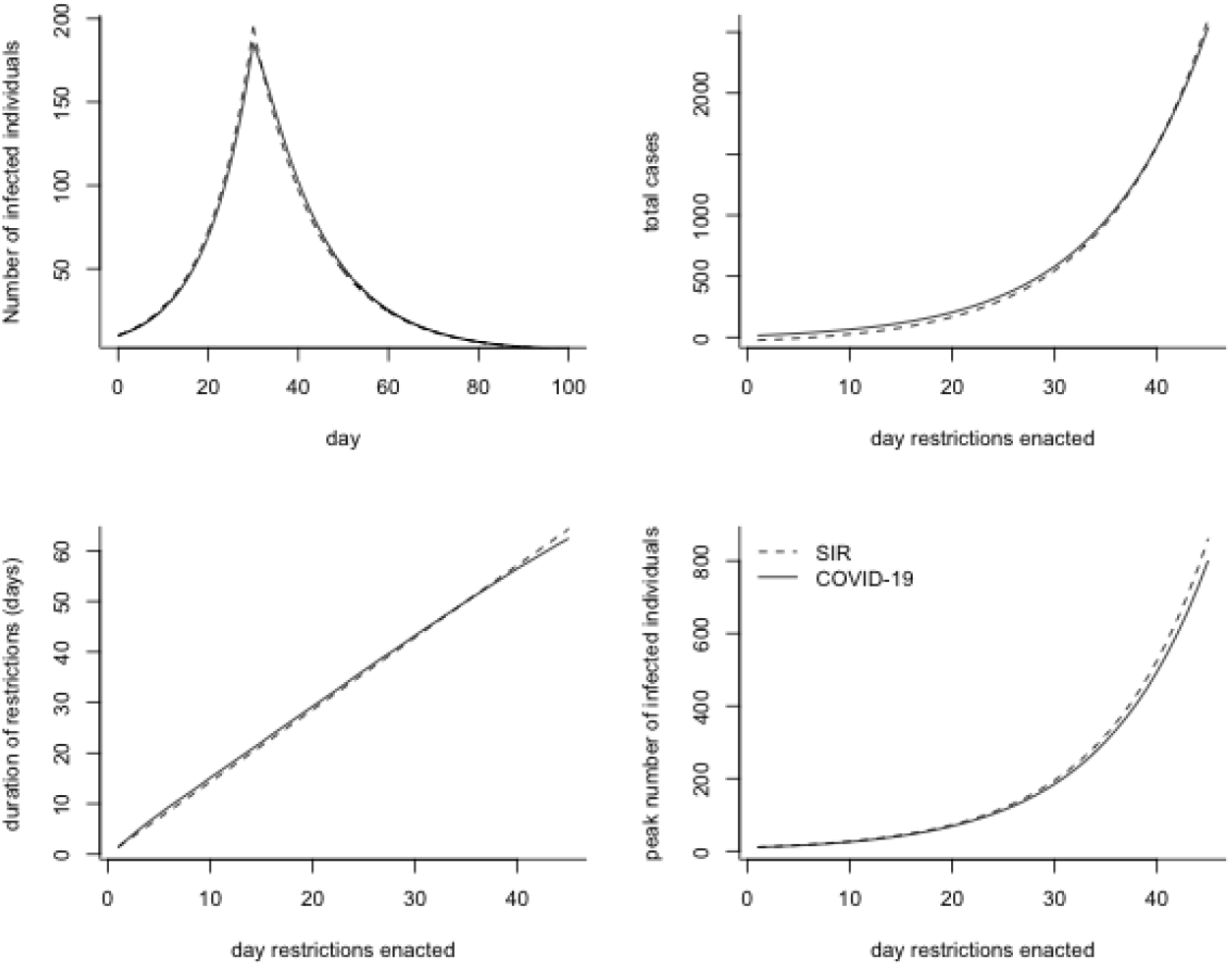
The dynamics and predictions of the linear SIR model are in close agreement with the nonlinear COVID-19 model. We find a close agreement between the linear SIR model (dashed lines; Equation 1a) and the COVID-19 model (solid lines; Equations 10) for the number of infected individuals over time (top left), the total number of cases during an outbreak (top right), the duration of restrictions (bottom left), and the peak number of infected individuals (bottom right). For the COVID-19 model, the number of infected individuals is calculated as *I*_*P*_(*t*) + *I*_*C*_(*t*) + *I*_*A*_(*t*) + *L*(*t*). All parameters values are listed in Table 4.

We found a close agreement between the predictions of the linear SIR model and these same quantities calculated from data (Table 1), suggesting that the linear SIR model can effectively predict the necessary duration of restrictions.

## 4 Comparison of the linear SIR model to a nonlinear COVID-19 model

The linear SIR model (Equations 1) assumes that the number of susceptible individuals is not changing, and does not distinguish between different types of infected individuals, while deterministic mathematical models specifically developed for COVID-19 typically include these features. In this section, we examine the agreement between the predictions of the linear SIR model and a nonlinear COVID-19 model with different types of infected individuals, where the comparison between these models is illustrated in Figure 4. The nonlinear COVID-19 model that we consider is based on [14], but without any age or spatial structure. The COVID-19 model equations are as follows:

**Fig. 4.**
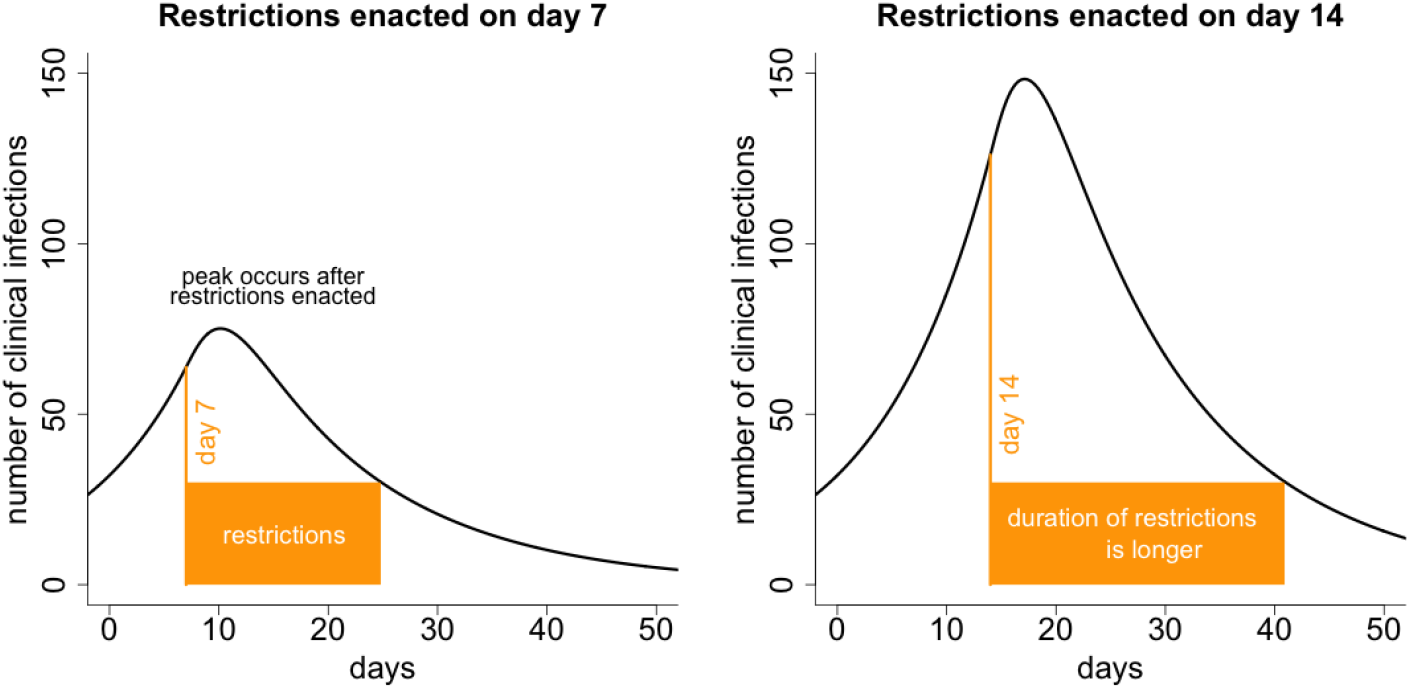
Delayed action results in a longer duration of COVID-19 restrictions. We calculated the number of active clinical infections, *I*_*C*_(*t*), (black line) for the COVID-19 model (Equations 10) with restrictions enacted on either day 7 (left) or day 14 (right) and remaining in place until there are 30 or fewer active clinical cases of COVID-19 (orange boxes). Delaying action results in a longer duration of restrictions, because a constant decay rate beginning from a higher peak will require longer to reduce the number of cases to the same level. The peak in the number of clinical cases occurs after the enactment of the restrictions because some individuals that were infected prior to the restrictions have yet to show symptoms. Elsewhere in this chapter (Equation 5 and Figure 4), we have shown that delaying restrictions results in a longer period of restrictions generally, not just for restrictions at 7 and 14 days as illustrated here. All parameter values are listed in Table 4.

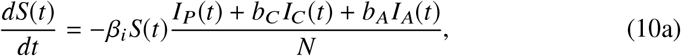

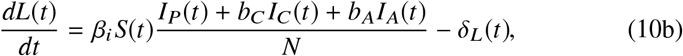

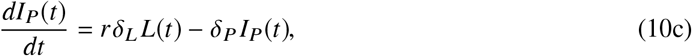

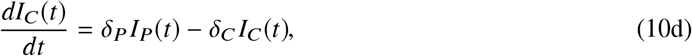

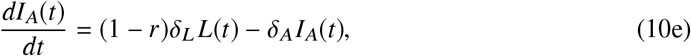

where *S (t)* is the number of susceptible individuals, *L*(*t*) is the number of latently infected individuals (infected, but not yet infectious), *I*_*P*_(*t*) is the number of individuals that have pre-clinical infections (infectious, but yet to be symptomatic), *I*_*C*_(*t*) is the number of individuals that have clinical infections (infectious and symptomatic), and *I*_*A*_(*t*) is the number of individuals that have asymptomatic infections (infectious, but without symptoms). All model parameters are defined and estimated in Table 4.

For these more complex COVID-19 mathematical models, analytic solutions such as the results in the *Linear SIR model* section are not possible. This model (Equations 10) is a variation on a class of SLIAR models previously studied for influenza and SARS-CoV-2 [3, 5]. Several relevant results for this class of models, including general formulae for the basic reproduction number and the final size of the epidemic were established earlier [4].

Many models for SARS and influenza pandemics, including those for the current COVID-19 pandemic, fall into the following general form:

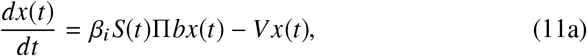

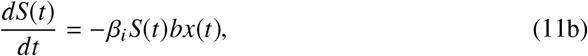

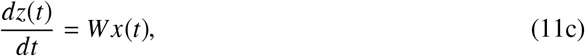

where *x*(*t*) is a vector of numbers of infected individuals in each stage, *S*(*t*) is again the dynamically changing number of susceptible individuals, and *z*(*t*) is a vector of numbers of individuals recovered through various routes of progression (typically either recovered from asymptomatic infection, recovered with only mild symptoms, recovered following hospitalization, or deceased). The parameter *β*_*i*_ is as defined previously, the vectors Π and *b* indicate the initial stage(s) of infection and relative transmission rates of the infectious stages, respectively, such that Π*b* is a rank one matrix. The matrices *V* and *W* contain rates of progression through and out of the stages of infection and recovery. This is a simplification of the Arino et al model which included a structured susceptible population [4]. The COVID-19 model (Equations 10) is the special case of the general SLIAR model with *x*(*t*) = *L*(*t*), *I*_*P*_ *(t), I*_*C*_(*t*), *I*_*A*_(*t*), Π = (1, 0, 0, 0) ^*T*^, *b* = (0, 1, *b*_*C*_, *b*_*A*_), and *V* a 4× 4 matrix containing the remaining parameters. The total numbers of observed cases, *C*(*t*), can be included by setting *z*(*t*) = *C*(*t*)and *W* =(0, *δ*_*P*_, 0, 0).

Following the same analysis as we have done with the linear SIR model, we extend the Equations 11a to include importation of infected individuals at rate *m*_*i*_ and linearize assuming *S*(*t*) remains near *S*_0_:

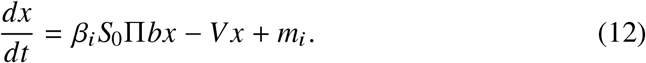

Although a simple integration leads to solutions for *C*(*t*_*1*_) and *C*(*t*_2_), the resulting transcendental relations do not obviously lead to simple expressions for the duration of restrictions or the total number of cases as with the linear SIR model. Hence, we instead proceed to a numerical comparison of the simple linear SIR model (Equations 1) and the COVID-19 model (Equations 10).

We assumed that the initial number of individuals in each infected state for the COVID-19 model (Equations 10) was close to the right eigenvector (Table 4). We numerically solved Equations 10 in R using the deSolve package [16]. We compared the number of infected individuals under each model formulation (*I*(*t*) for the linear SIR model and *L*(*t*) + *I*_*P*_(*t*)+ *I*_*C*_(*t*) + *I*_*A*_(*t*) for the COVID-19 model). Figure 4 shows a close agreement between the dynamics of the linear SIR model (Equation 1a) and the nonlinear COVID-19 model (Equations 10). All data and code for this Chapter are archived at https://github.com/ahurford/reescalation-chapter.

## 5 Discussion

We derive some simple equations to calculate the required duration of restrictions necessary to meet a target number of active infections (Equation 4) and the total number of cases in an outbreak (Equation 8), and while the model itself (Equation 1a) lacks complexity, when parameterized properly (Table 4), the dynamics are nearly identical to those of more complex nonlinear coupled ordinary differential equations (Equations 10, i.e. [14]). We also show that it is simple to estimate the parameters for the linear SIR model from epidemic data (Figure 3), which then allows for the duration of restrictions and the total number of cases to be predicted using Equations 4 and 8. From the expression for the necessary duration of restrictions (Equation 4), we are able to show that restrictions always need to be in place longer when actions are delayed (Equation 5).

### 5.1 Public health implications

Whether public health measures will be enacted to mitigate the spread of COVID-19 is a decision that public health officials should make prior to the discovery of exponentially growing case numbers in the local community. Regarding SARS-CoV-2, the novel coronavirus, whereby the majority of the population is susceptible to infection, waiting to enact the restrictions only delays the inevitable, whereby restrictions with undesirable impacts will still have to occur, just at a later date. However, while action is delayed, the number of cases will continue to increase exponentially, and an unintuitive consequence of delayed restrictions, is that the restrictions, and their undesirable impacts, will need to be in place longer, to reduce the number of cases to the same level (Figure 4).

While delayed restrictions might be justified to protect the economy and keep children in school, this is short sighted: delayed action will yield these benefits *now*, while ultimately these impacts will still be felt, just later and for longer. The only reasons to delay restrictions are if (1) a vaccine is likely in the near future, (2) conditions are likely to change in the near future, for example, a school break or a low tourism season, (3) there was never any intention to implement any restrictions, or (4)infection prevalence in the community is very low, such that exponential growth has not been established, and the outbreak may go extinct without any intervention. Frequently, the question has been posed ‘what should be a trigger for lockdowns?’, where the trigger is in terms of the number of active cases [13]. We suggest that, providing that exponential growth is established, there is no benefit to delaying action. Such early action does not prioritize epidemiology over other considerations, because early action also enables the restrictions to be lifted after a shorter time, allowing for unrestricted economic, social, and education activities to resume sooner, and potentially allowing for a longer period of unrestricted activities until restrictions need to be enacted again.

Finally, while the effective reproduction number, *R*_*t*_, has become a popular method for communicating epidemic trends, this metric has limitations, including that *R*_*t*_ is a lagged measure since the number of secondary infections generated per infected person can only be known after that person recovers, or by making assumptions via the method of nowcasting [2]. By contrast, the exponential growth rates, *λ*_1_, and *λ*_2_ are instantaneous measures, and can be easily communicated as doubling times or halving times. As an alternative to *R*_*t*_, we suggest estimating *λ*_1_, *λ*_2_ and *m*_1_, as shown in Figure 3, using Equation 1a to predict the future time course of the epidemic, and using Equation 4 to communicate the likely duration of restrictions shortly after they have been enacted.

Our key message is succinctly summarized by the phrase ‘Don’t wait, re-escalate’ as coined by Christina Bancej of the Public Health Agency of Canada. We find that delaying re-escalation of restrictions to prevent the spread of COVID-19 results in not only more infections, but also longer periods of restrictions. As such, we recommend not waiting to enact restrictions.

## Data Availability

All data and code are available at the link provided.

https://github.com/ahurford/reescalation-chapter

## Notes

### Competing Interest Statement

The authors have declared no competing interest.

### Funding Statement

AH and JW are funded by each by NSERC Discovery Grants and the AARMS Centre for Disease Modelling.

### Summary of Updates

Minor typos corrected.

